# The influence of APOE status on cognitive decline

**DOI:** 10.1101/2023.05.23.23290400

**Authors:** Cassandra Morrison, Michael D. Oliver, Virginia Berry, Farooq Kamal, Mahsa Dadar

## Abstract

**INTRODUCTION:** Apolipoprotein (APOE) ε4 and subjective cognitive decline (SCD) increase risk of Alzheimer’s disease. However, few studies have examined the relationship between SCD and APOE, especially using longitudinal data. The current study examined whether APOE is associated with the rate of cognitive change in SCD.

**METHODS:** Linear mixed effects models examined the relationship between APOE status and cognitive change in older adults with SCD (SCD), normal controls (NC), and people with mild cognitive impairment (MCI).

**RESULTS:** The presence of at least one ε2 allele in SCD and MCI results in cognitive change rates similar to a NC with the ε3ε3 genotype. Older SCD-ε4 individuals exhibited increased cognitive decline compared to all groups, including NC-ε4 and MCI-ε4.

**DISCUSSION:** People with SCD with at least one ε4 allele will experience increased cognitive decline compared to cognitively healthy older adults and people with MCI. These findings have important implications for treatments and interventions.

## 1. Introduction

Affecting nearly 55 million people worldwide, Alzheimer’s disease (AD) is by far the most common cause of dementia^1^. Functionally, AD is characterized by a progressive decline in memory and other cognitive abilities, resulting in the inability to perform normal activities of daily living^2^. People who exhibit deficits in cognitive functioning but maintain the ability to perform daily life activities independently, may be at a midpoint between healthy cognitive aging and dementia and are categorized as having mild cognitive impairment (MCI)^2,3^. The structural brain changes that result in these functional declines have been suggested to occur nearly 20 years prior to the onset of clinical symptoms^4^. This knowledge has led to a plethora of research targeting factors associated with increased risk and subsequent development of AD.

Several risk factors have been identified to be associated with AD. One of the most significant risk factors is a genetic factor from the variant of the Apolipoprotein E (APOE) gene. Those who have an APOE ε4 allele have an increased risk of AD^5^. Although the presence of an ε4 allele does not necessarily imply that one will develop AD, studies have revealed that heterozygous ε4 carriers have a 3-4 times higher risk, and homozygous ε4 carriers have an 8-12 times increased risk for AD development compared to non-carriers^6^. Research has also shown that individuals who are ε4 positive have significantly different trajectories in the clinical manifestation and pathogenesis of disease. For example, ε4 carriers exhibit an earlier age of AD onset^5^ and increased rates of cognitive decline^7^ compared to those who are not ε4 carriers. On the other hand, the ε2 allele has been shown to offer protective effects against risk of AD^8^, while ε3 does not offer any protective or detrimental effects towards AD^9^.

Another factor that increases risk of MCI^10^, AD development^11,12^, and cognitive decline^11^ is subjective cognitive decline (SCD). SCD is self-reported cognitive decline in older adults with no objective evidence of cognitive decline^11^, and is now thought to be the earliest stage of AD. Some research has suggested that compared to cognitively healthy older adults without SCD, those with SCD show higher frequencies of APOE ε4 positivity^13^. However, a systematic review observed that ε4 frequency does not differ between cognitively healthy older adults with and without SCD^14^. This review observed that both APOE positivity and SCD provide individual and multiplicative risks towards cognitive decline. While they indicate that SCD status and APOE ε4 positivity increase risk of developing objective cognitive impairment, they note that a limited understanding of the relationship between SCD and APOE status still remains, especially given the paucity of studies that have employed longitudinal cohorts^14^.

This paper was designed to expand on the current understanding of the relationship between preclinical AD and APOE status. To do so, we compared different APOE profiles in cognitively healthy older adults with and without SCD and people with MCI. This study design allowed us to examine the rate of cognitive change in people with SCD and whether APOE status influences the rate of change in this subset compared to cognitively healthy older adults without SCD and people with MCI. The main goal of this study was to determine if APOE status would influence the rate of cognitive change in people with SCD and MCI. That is, do people with SCD and MCI who have an ε2 allele have similar rates of cognitive decline to cognitively healthy older adults, and do people with SCD who have at least one ε4 allele have increased rates of cognitive decline compared to cognitively older adults and people with MCI with at least one ε4 allele.

## 2. Methods

### 2.1 Participants

Data used in preparation of this article were obtained from the RADC Research Resource Sharing Hub (www.radc.rush.edu). Participants provided informed written consent to participate in one of three cohort studies on aging and dementia: 1) Minority Aging Research Study (MARS)^15^, 2) Rush Alzheimer’s Disease Center African American Clinical Core (RADC AA Core)^16^, or 3) the Rush Memory and Aging Project (MAP)^17^.

Participant inclusion criteria for this specific study were as follows: 1) either cognitively healthy status or MCI status at baseline, 2) had at least two cognitive assessments, 3) were at least 55 years of age at baseline, and 4) had APOE genotyping completed. For cognitively healthy older adults to be included they must have also completed the questionnaire assessing memory complaints. A clinical diagnosis of cognitive status was completed using a three-stage process including computer scoring of cognitive tests, clinical judgment by a neuropsychologist, and diagnostic classification by a clinician based on criteria of the joint working group of the National Institute on Aging and the Alzheimer’s Association (NIA-AA)^18^.

Participants were divided into one of three diagnostic groups [cognitively normal (NC), subjective cognitive decline (SCD), and mild cognitive impairment (MCI)] and one of three APOE profiles [ε2 = ε2ε2 or ε2ε3; ε3= ε3ε3; or ε4 = ε4ε4 or ε4ε3]. To ensure group differences in follow-up duration did not impact the results, participants from each diagnostic group were also matched based on follow-up year. A total of 3494 older adults (NC = 1990, SCD = 775, MCI = 729), with a mean maximum follow-up of 9.09 years (and a total of 33722 follow-up timepoints) were included in this study. Participant demographic information by group is presented in Table 1.

**Table 1:** Participant demographic information by group

|  | NC | SCD | MCI |
| --- | --- | --- | --- |
| Number of participants | 1990 | 775 | 729 |
| ε2 group | 288 (14.4%) | 112 (14.5%) | 93 (12%) |
| ε3 group | 1267 (63.6%)* | 477 (61.5%) | 406 (54%) |
| ε4 group | 435 (22%)* | 186 (24%) | 230 (31%) |
| Age (years) | 75.3 ± 7.2* | 76.5 ± 7.42* | 78.9 ± 7.54 |
| Sex (females) | 1568 (79%) | 584 (75%) | 535 (72%) |
| Education (years) | 15.9 ± 3.9 | 15.7 ± 3.8 | 15.6 ± 3.6 |
| Follow-up timepoints | 19783 | 7466 | 6454 |
| Mean maximum follow-up (years) | 9.4 ± 5.5 | 9.1 ± 5.3 | 8.3 ± 4.7 |
| Medium follow-up (years) | 8 | 8 | 7 |

Notes: numbers are presented as total (percentage of group) or mean ± standard deviation. NC = cognitively normal controls; SCD = subjective cognitive decline; MCI = mild cognitive impairment. NC was younger than MCI. SCD was younger than MCI. MCI had increased ε4 and decreased ε3 frequency compared to NC.

Subjective cognitive decline was defined based on two questions examining memory complaints. Participants were asked, “About how often do you have trouble remembering things?”and “Compared to 10 years ago, would you say that your memory is much worse, a little worse, the same, a little better, or much better?”. Both questions were scored using a scale of 1 to 5 with 1 being never/much better and 5 being often/worse. Following past research and the Rush recommendations, if the participants had a composite score of 8-10 on these two questions they were classified as having memory complaints ^19^; reported as subjective cognitive decline (SCD+) in this paper.

### 2.2 Cognitive Scores

A neurological battery comprising 19 cognitive assessments was administered to all participants annually^15^. Five cognitive domains were assessed through the selected 19 tests: episodic memory, semantic memory, working memory, visuospatial ability, and perceptual speed. Episodic memory was assessed through scores from Word List Memory, Word List Recall, Word List Recognition, and immediate and delayed recall scores of both Story A on Logical Memory and the East Boston Story. Semantic memory was assessed through performance on Verbal Fluency, and on both a 15-item reading test, and a version of the Boston Naming Test. The working memory domain was measured through performance on Digit ordering, as well as Digit Span Forwards and Backwards. Visuospatial ability was assessed through performance on a 16-item version of the Progressive Matrices and a 15-item version of Judgement and Line Orientation. Perceptual speed was measured through performance on Number Comparison, two indices from a modified version of the Stroop Neuropsychological Screening Test, and the Symbol Digit Modalities Test. Raw scores on each of the individual tests were converted to z-scores using the baseline mean (SD); and the z-scores of the tests from each domain were then averaged. An individual’s standard performance across all 19 of these tests was averaged to create a measure of global cognitive function^20^. More information for the specific tests used for each category can be obtained from https://www.radc.rush.edu/.

### 2.3 Statistical Analysis

Analyses were performed using ‘R’ software version 4.0.5. Independent sample t-tests and chi-square analyses were completed on demographic information with multiple comparisons corrected for using Bonferroni correction. Linear mixed effects models were used to investigate the association between each group (NCε2, NCε3, NCε4, SCDε2, SCDε3, SCDε4, MCIε2, MCIε3, and MCIε4) and rate of cognitive change for each cognitive domain (global, episodic memory, semantic memory, perceptual speed, working memory, and perceptual orientation). All continuous values (except follow-up year) were z-scored within the population prior to analyses. Follow-up year was not z-scored to allow for the calculation of annual rate of change. All results were corrected for multiple comparisons using false discovery rate (FDR) of 0.05, p-values are reported as raw values with significance then determined by FDR correction^21^.

The first analysis was completed within each diagnostic category individually (NC, SCD, and MCI separately) to determine within group effects of APOE status. The interaction of interest was TimeFromBaseline:Group to determine if rate of cognitive decline differed within each APOE group (i.e., ε2, ε3, ε4) in each diagnostic category.

The second analysis used the same model but included SCD, NC, and MCI in the same model. This analysis was used to allow for comparison of the impact of APOE status on cognitive decline in people with SCD to that of the other diagnostic groups. The interaction of interest was TimeFromBaseline:Group to estimate the annual rate of cognitive decline in each group and determine if cognitive change over time differed for SCD groups compared to NC and MCI.

Participant ID was included as a categorical random effect to account for repeated measures of the same participant. Baseline age (Age_bl), sex, years of education, TimeFromBaseline, and Group were included as covariates in all models.

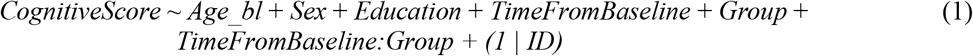

Annual rate of change was calculated for each group based on the estimated slope for each group and cognitive domain. Cognitive status at 20 years of follow up was also calculated using the model estimates [i.e., intercept + (slope*year) with the value for year being 20].

## 3. Results

### 3.1 Demographics

Baseline demographic information differed between the groups in terms of age and APOE status. Normal controls were younger than SCD (*t* = 4.05, *p*<.001) and MCI (*t*=11.67, *p*<.001), and SCD was younger than MCI (*t* = 6.51, *p*<.001). MCI participants had increased frequencies of ε4 allele (x^2^ = 11.71, *p*<.001) and decreased ε3 allele frequencies (x^2^ = 14.71, *p*<.001) compared to NC. No other differences remained significant after correction for multiple comparisons.

### 3.2 Within group impact of APOE on cognitive decline

Table 1 presents the TimeFromBaseline:Group interaction estimates from the linear mixed effects model examining the influence of APOE status on rate of cognitive decline within each group (NC, SCD, and MCI) individually.

For the NC, people with ε2 had a slower rate of cognitive decline compared to those with ε4 and ε3 for all cognitive domains [*t* belongs to (−10.50 – -2.34), *p*<.05] except for perceptual orientation in those with an ε3 (*p*>.05). Furthermore, people with ε3 had a slower rate of cognitive decline compared to ε4 in all domains [*t* belongs to (−8.33 – -3.29), *p*<.005]. For the SCD group, people with ε2 had a slower rate of cognitive decline than ε3 in global cognition, semantic memory, and working memory [*t* belongs to (−2.93 – -3.99), *p*<.005], and a slower rate of change than people with ε4 in global cognition, episodic memory, perceptual orientation, semantic memory, and working memory [*t* belongs to (−8.91 – -2.49), *p*<.05]. For the MCI group, people with ε2 did not have a rate of cognitive decline that differed from those with an ε3 in any cognitive domain. Those with an ε2 and ε3 had a lower rate of cognitive decline than ε4 in global cognition, episodic memory, and semantic memory [*t* belongs to (−4.35 – -2.29), *p*<.05].

### 3.3 Between group impact of APOE status on cognitive decline

Table 2 presents the TimeFromBaseline:Group interaction examining between group effects on rate of change for each cognitive domain. Table 3 presents the annual rate of change for each group by APOE status.

**Table 2:** Linear mixed effects model outputs for analysis 1 showing the TimeFromBaseline:Group for each diagnostic category separately.

| Within Group Effects | Global | Episodic Memory | Perceptual Orientation | Perceptual Speed | Semantic Memory | Working Memory |
| --- | --- | --- | --- | --- | --- | --- |
| NCε2:NCε3 | $\beta = -0.07$, SE= 0.01, $t = -5.20$, $p < .001$ | $\beta = -0.06$, SE= 0.01, $t = -3.92$, $p < .001$ | $\beta = -0.01$, SE= 0.01, $t = -0.43$, $p = .66$ | $\beta = -0.04$, SE= 0.01, $t = -3.48$, $p < .001$ | $\beta = -0.05$, SE= 0.01, $t = -3.41$, $p < .001$ | $\beta = -0.03$, SE= 0.01, $t = -2.34$, $p = .019$ |
| NCε2:NCε4 | $\beta = -0.07$, SE= 0.01, $t = -10.50$, $p < .001$ | $\beta = -0.12$, SE= 0.02, $t = -7.89$, $p < .001$ | $\beta = -0.05$, SE= 0.02, $t = -2.75$, $p = .005$ | $\beta = -0.09$, SE= 0.01, $t = -6.19$, $p < .001$ | $\beta = -0.12$, SE= 0.02, $t = -7.84$, $p < .001$ | $\beta = -0.09$, SE= 0.02, $t = -5.78$, $p < .001$ |
| NCε3:NCε4 | $\beta = -0.09$, SE= 0.01, $t = -8.30$, $p < .001$ | $\beta = -0.07$, SE= 0.01, $t = -6.19$, $p < .001$ | $\beta = -0.04$, SE= 0.01, $t = -3.29$, $p = .001$ | $\beta = -0.05$, SE= 0.01, $t = -4.40$, $p = .001$ | $\beta = -0.07$, SE= 0.01, $t = -6.77$, $p < .001$ | $\beta = -0.06$, SE= 0.01, $t = -5.18$, $p < .001$ |
| SCDε2:SCDε3 | $\beta = -0.06$, SE= 0.02, $t = -2.93$, $p = .002$ | $\beta = -0.02$, SE= 0.02, $t = -0.81$, $p = .42$ | $\beta = -0.03$, SE= 0.02, $t = -1.27$, $p = .20$ | $\beta = -0.02$, SE= 0.02, $t = -2.15$, $p = .031$ | $\beta = -0.09$, SE= 0.02, $t = -3.99$, $p < .001$ | $\beta = -0.07$, SE= 0.02, $t = -3.03$, $p = .001$ |
| SCDε2:SCDε4 | $\beta = -0.24$, SE= 0.03, $t = -8.91$, $p < .001$ | $\beta = -0.18$, SE= 0.03, $t = -6.50$, $p < .001$ | $\beta = -0.07$, SE= 0.02, $t = -2.49$, $p = .013$ | $\beta = -0.05$, SE= 0.02, $t = -1.01$, $p = .31$ | $\beta = -0.21$, SE= 0.03, $t = -7.65$, $p < .001$ | $\beta = -0.20$, SE= 0.03, $t = -7.17$, $p < .001$ |
| SCDε3:SCDε4 | $\beta = -0.17$, SE= 0.02, $t = -8.30$, $p < .001$ | $\beta = -0.16$, SE= 0.02, $t = -7.48$, $p < .001$ | $\beta = -0.04$, SE= 0.02, $t = -1.84$, $p = .07$ | $\beta = -0.03$, SE= 0.02, $t = -1.68$, $p = .09$ | $\beta = -0.12$, SE= 0.02, $t = -5.55$, $p < .001$ | $\beta = -0.13$, SE= 0.02, $t = -5.94$, $p < .001$ |
| MCIε2:MCIε3 | $\beta = -0.05$, SE= 0.04, $t = -1.38$, $p = .17$ | $\beta = -0.01$, SE= 0.04, $t = -0.27$, $p = .79$ | $\beta = -0.06$, SE= 0.04, $t = -1.48$, $p = .14$ | $\beta = -0.01$, SE= 0.03, $t = -0.11$, $p = .91$ | $\beta = -0.04$, SE= 0.04, $t = -1.14$, $p = .25$ | $\beta = -0.06$, SE= 0.04, $t = -1.64$, $p = .10$ |
| MCIε2:MCIε4 | $\beta = -0.12$, SE= 0.04, $t = -3.06$, $p = .001$ | $\beta = -0.09$, SE= 0.04, $t = -2.29$, $p = .022$ | $\beta = -0.07$, SE= 0.04, $t = -1.67$, $p = .09$ | $\beta = 0.01$, SE= 0.04, $t = 0.29$, $p = .77$ | $\beta = -0.16$, SE= 0.04, $t = -3.90$, $p < .001$ | $\beta = -0.09$, SE= 0.04, $t = -2.19$, $p = .028$ |
| MCIε3:MCIε4 | $\beta = -0.07$, SE= 0.02, $t = -2.69$, $p = .007$ | $\beta = -0.05$, SE= 0.03, $t = -3.15$, $p = .001$ | $\beta = -0.01$, SE= 0.03, $t = -0.39$, $p = .70$ | $\beta = 0.01$, SE= 0.02, $t = 0.62$, $p = .54$ | $\beta = -0.11$, SE= 0.03, $t = -4.35$, $p < .001$ | $\beta = -0.02$, SE= 0.02, $t = -0.97$, $p = .33$ |

NC = cognitively healthy older adults without subjective cognitive decline. SCD = cognitively healthy older adults with subjective cognitive decline. MCI = mild cognitive impairment. ε2 = someone with either ε2 ε3 or ε2 ε2. ε3 = someone with ε3 ε3. ε4 = someone with ε4 ε3 or ε4 ε4. Bold values are those that remained significant after correction for multiple comparisons.

**Table 3:** Linear mixed effects model outputs for analysis 2 showing the TimeFromBaseline:Group interaction examining between group effects.

| Between Group Effects | Global | Episodic Memory | Perceptual Orientation | Perceptual Speed | Semantic Memory | Working Memory |
| --- | --- | --- | --- | --- | --- | --- |
| NCε4:SCDε2 | **$\beta = -0.02$, SE<0.01, $t = -3.85$, $p < .001$** | **$\beta = -0.01$, SE<0.01, $t = -2.22$, $p = .025$** | $\beta = -0.01$, SE<0.01, $t = -1.95$, $p = .052$ | **$\beta = -0.01$, SE<0.01, $t = -2.75$, $p = .005$** | **$\beta = -0.02$, SE<0.01, $t = -4.02$, $p < .001$** | **$\beta = -0.01$, SE<0.01, $t = -3.06$, $p = .022$** |
| NCε3:SCDε2 | $\beta < 0.01$, SE<0.01, $t = 0.25$, $p = .71$ | $\beta < 0.01$, SE<0.01, $t = 0.89$, $p = .37$ | $\beta < -0.01$, SE<0.01, $t = -0.31$, $p = .75$ | $\beta < -0.01$, SE<0.01, $t = -0.53$, $p = .60$ | $\beta < -0.01$, SE<0.01, $t = -0.71$, $p = .48$ | $\beta < -0.01$, SE<0.01, $t = -0.40$, $p = .69$ |
| NCε4:MCIε2 | **$\beta = -0.02$, SE<0.01, $t = -3.65$, $p < .001$** | **$\beta = -0.03$, SE<0.01, $t = -4.08$, $p < .001$** | **$\beta = -0.03$, SE<0.01, $t = -4.72$, $p < .001$** | **$\beta = -0.03$, SE<0.01, $t = -3.01$, $p = .003$** | $\beta = -0.01$, SE<0.01, $t = -2.03$, $p = .043$ | **$\beta = -0.02$, SE<0.01, $t = -2.45$, $p = .014$** |
| NCε3:MCIε2 | $\beta < -0.01$, SE<0.01, $t = -0.69$, $p = .49$ | $\beta < -0.01$, SE<0.01, $t = -1.93$, $p = .053$ | **$\beta = -0.02$, SE<0.01, $t = -3.65$, $p < .001$** | $\beta < -0.01$, SE<0.01, $t = -1.42$, $p = .16$ | $\beta < -0.01$, SE<0.01, $t = -0.45$, $p = .65$ | $\beta < -0.01$, SE<0.01, $t = -0.52$, $p = .60$ |
| NCε4:SCDε4 | **$\beta = 0.03$, SE<0.01, $t = 7.74$, $p < .001$** | **$\beta = 0.03$, SE<0.01, $t = 6.26$, $p < .001$** | $\beta < 0.01$, SE<0.01, $t = 1.13$, $p = .29$ | $\beta < -0.01$, SE<0.01, $t = -0.23$, $p = .82$ | **$\beta = 0.02$, SE= 0.02, $t = 5.54$, $p < .001$** | **$\beta = 0.03$, SE= 0.02, $t = 6.00$, $p < .001$** |
| MCIε4:SCDε4 | **$\beta = 0.03$, SE<0.01, $t = 6.16$, $p < .001$** | **$\beta = 0.03$, SE<0.01, $t = 6.74$, $p < .001$** | **$\beta = 0.02$, SE<0.01, $t = 4.09$, $p < .001$** | **$\beta = 0.02$, SE<0.01, $t = 3.98$, $p < .001$** | $\beta < 0.01$, SE<0.01, $t = 0.89$, $p = .38$ | **$\beta = 0.02$, SE= 0.02, $t = 4.82$, $p < .001$** |

NC = cognitively healthy older adults without subjective cognitive decline. SCD = cognitively healthy older adults with subjective cognitive decline. MCI = mild cognitive impairment. ε2 = someone with either ε2 ε3 or ε2 ε2. ε3 = someone with ε3 ε3. ε4 = someone with ε4 ε3 or ε4 ε4. Bold values are those that remained significant after correction for multiple comparisons.

**Table 4:** Annual rate of change by Group and cognition at 20 years

| Group | Global |  | Episodic Memory |  | Perceptual Orientation |  | Perceptual Speed |  | Semantic Memory |  | Working Memory |  |
| --- | --- | --- | --- | --- | --- | --- | --- | --- | --- | --- | --- | --- |
|  | Annual | 20 years | Annual | 20 years | Annual | 20 years | Annual | 20 years | Annual | 20 years | Annual | 20 years |
| NCε2 | -0.037 | -0.34 | -0.020 | -0.01 | -0.022 | -0.33 | -0.051 | -0.57 | -0.030 | -0.20 | -0.028 | -0.26 |
| NCε3 | -0.050 | -0.60 | -0.030 | -0.20 | -0.023 | -0.32 | -0.060 | -0.71 | -0.050 | -0.55 | -0.035 | -0.45 |
| NCε4 | -0.067 | -0.94 | -0.044 | -0.58 | -0.031 | -0.49 | -0.069 | -0.93 | -0.066 | -0.96 | -0.047 | -0.69 |
| SCDε2 | -0.051 | -0.62 | -0.034 | -0.28 | -0.022 | -0.30 | -0.062 | -0.87 | -0.047 | -0.59 | -0.033 | -0.36 |
| SCDε3 | -0.064 | -0.93 | -0.038 | -0.51 | -0.028 | -0.44 | -0.062 | -0.89 | -0.066 | -0.97 | -0.047 | -0.64 |
| SCDε4 | -0.099 | -1.63 | -0.070 | -1.15 | -0.036 | -0.61 | -0.068 | -1.10 | -0.090 | -1.45 | -0.075 | -1.20 |
| MCIε2 | -0.054 | -0.98 | -0.019 | -0.42 | 0.00 | -0.55 | -0.052 | -0.92 | -0.053 | -1.16 | -0.032 | -0.84 |
| MCIε3 | -0.056 | -1.42 | -0.021 | -0.78 | -0.012 | -0.74 | -0.053 | -1.04 | -0.060 | -1.4 | -0.044 | -1.03 |
| MCIε4 | -0.070 | -2.00 | -0.037 | -1.47 | -0.014 | -0.83 | -0.050 | -1.25 | -0.085 | -2.05 | -0.049 | -1.23 |

SCD = Subjective cognitive decline. NC = Normal Controls. MCI = mild cognitive impairment. ε2 = someone with either ε2ε3 or ε2ε2. ε3 = someone with ε3ε3. ε4 = someone with ε4ε3 or ε4ε4. The annual rate of change reflects slope of the line in Figure 1 and 20 years represent the value of the line (i.e., cognitive score) at 20 years. The red shaded boxes represent the three groups with the largest annual rate of change and the blue shaded boxes represent the three groups with the lowest cognitive score at 20 years.

Older adults with SCD-ε2 had a slower rate of cognitive decline compared to NC-ε4 in all cognitive domains [*t* belongs to (−2.22 – -4.02), *p*<.05], except perceptual orientation in which their rate of change did not differ. Additionally, MCI-ε2 also had slower rates of cognitive decline compared to NC-ε4 in all cognitive domains [*t* belongs to (−2.45 – -4.72), *p*<.05], except semantic memory in which their rate of change did not differ. Both SCD-ε2 and MCI-ε2 did not differ from NC-ε3 in any cognitive domain (*p*<.05) except MCI-ε2 had a slower rate of change in perceptual orientation than NC-ε3 (*t= -*3.65, *p*<.001). That is, both people who have SCD and MCI who also have at least one ε2 (i.e., ε2ε2 or ε2ε3) have a lower annual rate of cognitive change compared to cognitively healthy older adults with at least one ε4 (i.e., ε4ε3 or ε4ε4) and similar rates of change to cognitively healthy older adults with the ε3ε3 genotype.

Older adults with SCD-ε4 exhibited increased rates of cognitive change compared to NC-ε4 in all cognitive domains [*t* belongs to (5.54 – 7.74), *p*<.001], except perceptual orientation and perceptual speed. Furthermore, this group (SCD-ε4) also exhibited increased rates of cognitive change compared to MCI-ε4 in all domains [*t* belongs to (3.98 – 6.74), *p*<.001] except semantic memory. That is, people in the SCD-ε4 had increased rates of annual change compared to all other ε4 groups except in a select few domains.

To ensure that differences were not being driven by differences in the individual cohorts we re-ran all analysis with study cohort as a random categorical effect + (1|study). The models produced similar results in terms of effect size and significance. These findings reflect an insignificant role of the study cohort on the current findings.

## 4. Discussion

While it is well-established that APOE status increases risk of AD^5,6^, few studies have examined how APOE status interacts with subjective cognitive decline to influence rate of cognitive change. To improve our understanding of how these two potentially independent risk factors (i.e., APOE and SCD status) interact to influence cognitive change, the current study examined the rate of cognitive change over a 28-year period in NCs, people with SCD, and people with MCI. The goal of the current study was to determine if SCD-ε2 and MCI-ε2 adults exhibit similar rates of cognitive decline to NCs, and if SCD-ε4 adults have increased rates of cognitive decline compared to NC-ε4 and MCI-ε4. In our sample of 3494 older adults, we observed that SCD-ε4 older adults had the largest rate of cognitive change compared to all other groups.

Importantly, this rate of change in SCD-ε4 was also steeper than both NC-ε4 and MCI-ε4 older adults. This study also revealed that SCD-ε2 and MCI-ε2 exhibit similar rates of cognitive change to NC-ε3 and less change than an NC-ε4. On the other hand, an ε4 increased rate of cognitive change, particularly in people with SCD, who had the steepest rate of change. Furthermore, consistent with previous findings, we did not observe differences in ε4 frequencies between NC and SCD^14^, supporting the hypothesis that SCD and APOE ε4 status independently influence longitudinal cognitive decline. We did, however, observe increased frequencies of ε4 and deceased frequencies of ε3 in people with MCI compared to NCs.

People who have SCD^22^ or MCI^23^ have been consistently observed to exhibit increased rates of cognitive change compared to NCs. However, in the current study, both SCD-ε2 and MCI-ε2, showed similar rates of change to NC-ε3 and less cognitive change than NC-ε4. This result is consistent with previous reports indicating that the presence of the ε2 allele offers protective effects against risk of AD^8^. These findings indicate that even when an older adult is further along the cognitive decline trajectory (i.e., SCD or MCI), having at least one ε2 allele can positively influence their rate of cognitive change. On the other hand, SCD-ε4 older adults had increased rates of cognitive change compared to both NC-ε4 and MCI-ε4, indicating that early in the decline process, APOE status is strongly associated with cognitive decline, more than in someone with MCI. These findings suggest that APOE status may be a strong risk factor for cognitive decline in people with SCD but is less strongly associated with decline once someone has cognitive impairment (i.e., MCI). This interpretation is further supported by the stronger within-group effects between APOE status and rate of cognitive change in SCD than in MCI.

Within SCD, people with an ε4 had steeper declines than ε2 in all domains except perceptual speed, and steeper declines than ε3 in all domains except perceptual orientation and speed. On the other hand, fewer effects (and smaller beta estimates) were observed for both the ε4:ε2 and ε4:ε3 differences within the MCI group. Cognitively healthy older adults with at least one ε4 (NC-ε4) also had increased rates of cognitive change compared to NC-ε3 and NC-ε2; however, as can be observed by the smaller beta estimates, the effects were less pronounced than in SCD. Overall, those with SCD-ε4 had the steepest rate of change indicating that in those who are cognitively unimpaired, a higher rate of cognitive impairment is associated with the ε4 allele^24^. These findings coincide with past research indicating that older adults with SCD who have an ε4 allele are at increased risk of being in the preclinical stage of AD^25^. Given that SCD may be the earliest pre-clinical stage of AD^11,12^, they are an important group to target for clinical trials and intervention strategies to reduce rate of cognitive decline before the irreversible AD-related pathology and neurodegeneration occurs. Therefore, observing that this groups’ cognition is most affected by APOE status has important implications for both research and clinical settings.

Given the multiplicative effects of SCD and APOE on AD risk^14^, and the fact that no cure or disease-modifying drug has become widely available for AD treatment, it is imperative to target both risk factors (i.e., SCD and APOE) early in order to better understand those at risk for the development of AD. AD pathology is thought to manifest up to two decades prior to observations of clinical symptoms^4^. Although objective decline may not be present during this stage, it is during this phase that SCD may occur^11^. Additionally, genetic testing is unlikely to occur during this stage unless there is a family history of AD. However, genetic testing may be especially important for those in the SCD stage. Our findings reveal that individuals with SCD may be most impacted by the presence of at least one ε4 allele, indicating that although ε4 increases risk for decline in all groups, the individuals most affected are the ones who are in the earliest stages of the disease process (i.e., those with SCD). Our findings are comparable with those of early-onset AD. For example, individuals living with early-onset AD experience greater cognitive change compared to late-onset AD^26,27^. It is thought that these early changes result from genetic mutations in amyloid precursor protein (APP), Presenilin-1 (PSEN1), or Presenilin-2 (PSEN2)^28^. As such, interventions have been proposed to target these genes in preclinical populations. Taken together, these results suggest that it is critical to also investigate APOE in preclinical stages of the disease as it is related to increased AD risk. Specifically, genetic testing for APOE, paired with regular cognitive assessments of memory complaints in cognitively normal adults may prove critical to the earliest possible treatment and/or intervention efforts. Given that APOE-specific interventions have been shown to be effective at mitigating cognitive decline in ε4 carriers^30^, capturing these individuals at the earliest possible stage (i.e., SCD) is important. This work can aid in cohort selection for clinical trials by identifying those at greater risk even before they experience objective decline. Our work suggests that clinical trials or interventions can be targeted toward those with APOE ε4 in individuals without objective decline, but especially so in those with subjective decline, to better understand the risk factors associated with AD, and develop treatments for AD prevention.

There are a few strengths and limitations of the study that should be noted. Firstly, this study leverages the RUSH dataset which is a large, diverse dataset, which has long follow-up durations. Long follow-up durations are important for this work because we are examining cognitive change either before (NCs) or early in the disease process (preclinical AD/SCD), and previous research has observed that the relationship between APOE ε4 status and cognitive decline is most pronounced after 7 years of follow-up^24^. Therefore, shorter follow-up durations may not capture the full relationship. A limitation of the current study is the comparison of APOE status to cognitive data only. Future research should examine the relationship between APOE status and structural brain changes in people with SCD. This design would help detect early brain biomarkers associated with disease progression in people who are ε4 positive.

Overall, our current findings show that the relationship between the APOE ε4 allele and cognition is strongest in cognitively unimpaired older adults with SCD. Thus, SCD-ε4 older adults experience an increased rate of cognitive decline which may put them at an increased risk for future AD because of reduced cognitive resilience. These findings can improve future research and clinical trials by targeting people in the preclinical AD phase (i.e., SCD) who also possess at least one APOE ε4 allele. Because these older adults (SCD-ε4) exhibit the steepest decline in functioning, using targeted interventions for these individuals would be the most beneficial to reduce cognitive decline.

**Figure 1:**
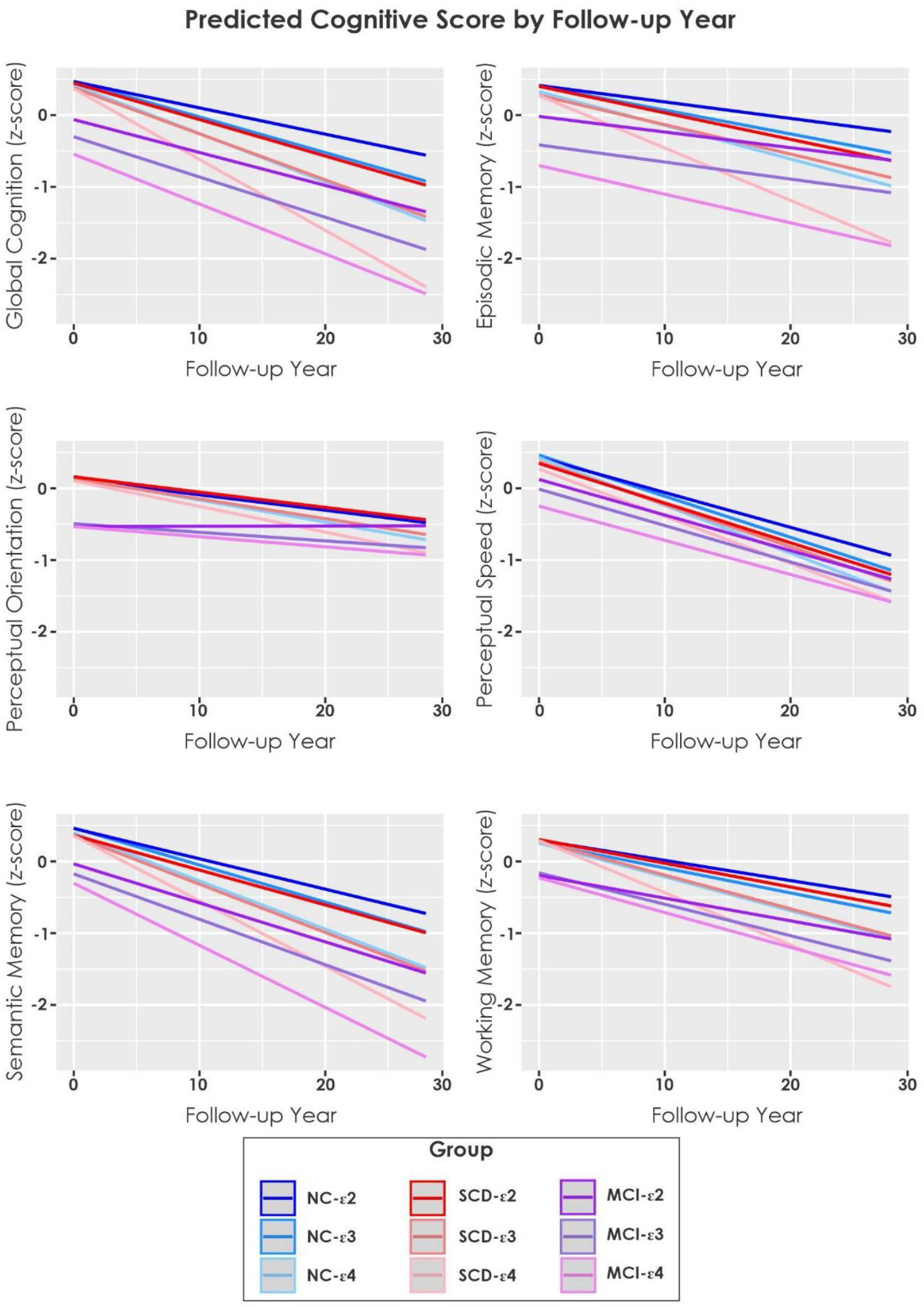
Estimated longitudinal cognitive change over time by group and cognitive domain. Notes: NC = cognitively normal controls. SCD = subjective cognitive decline. MCI = mild cognitive impairment.

## Data Availability

All data produced are available online at https://www.radc.rush.edu/.

## Acknowledgements

We want to acknowledge all the MARS, AA Core, and MAP participants. We are also grateful for the hard work from the staff and investigators at the Rush Alzheimer’s Disease Center. To obtain data from MARS, AA Core, and MAP for research use, please visit the RADC Research Resource Sharing Hub (www.radc.rush.edu).

## Financial Disclosures

Dr. Morrison is supported by a postdoctoral fellowship from Canadian Institutes of Health Research, Funding Reference Number: MFE-176608.

Dr. Kamal is supported by the Quebec Bioimaging Network and Fonds de Recherche du Québec (FRQS) postdoctoral scholarships.

Dr. Dadar reports receiving research funding from the Healthy Brains for Healthy Lives (HBHL), Alzheimer Society Research Program (ASRP), Natural Sciences and Engineering Research Council of Canada (NSERC), Fonds de Recherche du Québec (FRQS), and Douglas Research Centre (DRC).

The RADC/ RUSH cohort studies are supported by the National Institutes of Health, National Institute on Aging (R01 AG17917, R01 AG22018, P30 AG10161, and P30 AG72975).

## Conflict of Interest

The authors declare no competing interests.

## Notes

### Competing Interest Statement

The authors have declared no competing interest.

### Author Declarations

The study used (or will use) ONLY openly available human data that were originally located at: https://www.radc.rush.edu/.

